# Risk Assesment of nCOVID-19 Pandemic In India: A Mathematical Model And Simulation

**DOI:** 10.1101/2020.04.10.20060830

**Authors:** Swarnava Biswas, Moumita Mukherjee

**Affiliations:** Adamas University, Barasat - Barrackpore Rd, 24 Parganas North, Jagannathpur, Kolkata, West Bengal 700126

**Keywords:** nCOVID-19, Corona Virus, India, SEIR-Modelling, Epidemic Model

## Abstract

The entire world is now eventually locked down due to the outbreak of nCOVID-19 corona virus outbreak. The fast and relentless spread nCOVID-19 has basically segmented the populace only into merely into three classes, such as susceptible, infected and recovered compartments. Adapting the classical SEIR-type epidemic modelling framework, the direct person-to-person contact transmission is taken as the direct route of transmission of nCOVID-19 pandemic. In this research, the authors have developed a model of the nation-wide trends of the outburst of the nCOVID-19 infection using an SEIR Model. The SEIR dynamics are expressed using ordinary differential equations. The creators initially determined the parameters of the model from the accessible day by day information for Indian States dependent on around 35 days history of diseases, recuperations and deaths. The determined parameters have been amassed to extend future patterns for the Indian subcontinent, which is right now at a beginning time in the contamination cycle. The novelty of the study lies in the prediction of both the pessimistic and optimistic mathematical model based comprehensive analysis of nCOVID-19 infection spreading, for two different conditions: (a) if lockdown gets withdrawn and (b) if lockdown continues as a whole. If the complete lockdown in India is withdrawn on 14th April 2020, as a whole, then from the simulation, the authors have predicted that the infected population will flare-up to a large extent, suddenly, however, gradual or zone specific withdrawal would be more effective solution. This study also suggested some possible way-out to get rid of this situation by providing a trade-off between ‘ flattening of the curve” as well as “less economic turbulence. The projections are intended to provide a base/ action plan for the socio-economic counter measures to alleviate nCOVID-19.

## Introduction

Human coronaviruses were first recognized during the 1960s and enthusiasm for these infections progressed altogether in 2002 from the development of Severe Acute Respiratory Syndrome CoV (SARSCoV) which executed 774 individuals around the globe with a death pace of 9.5% ^[1-3]^. Later it infected the Middle East regions as the Middle East Respiratory Syndrome CoV (MERS-CoV), that killed 919 individuals with an exceptionally high mortality pace of 35% ^[4]^. In December 2019, it has returned in Wuhan, China and has spread to 199 nations killing 91783 people (World Health Organization 2020) to date and has been declared a pandemic by the World Health Organization. The Wuhan CoV viral genome sequencing shows less comparability with the previous detailed CoVs proposing a novel CoV strain (2019-nCoV), that has presently been named as Severe Acute Respiratory Syndrome CoV-2 (SARS-CoV-2) or COVID-19. 2019-nCoV is thought to arrive at people through creatures particularly bats and the specific method of transmission despite everything stay obscure ^[5]^. The death rate of this novel infection falls between 1-3% (Novel, C. P. E. R. E. 2020) and its transmission pace is extremely high looked at to its previous partners as it kills a greater number of individuals than the prior productive killer CoV strains.

2019-nCoV is a solitary stranded RNA infection that at first attacks the lung parenchyma and causes extreme interstitial aggravation of the lungs. The tainted people regularly have alveolar harm that could prompt intense respiratory misery disorder. The patient frequently experiences lymphocytopenia which frequently connects with the seriousness of the sickness. Intense respiratory pain disorder can bring about cytokine storm and non-administrative resistant reactions may in the end lead to various organ breakdowns. Most instances of 2019-nCoV diseases are related with pneumonia and injury in hepatic tissues ^[6]^.

According to a stochastic, around the world, air transportation arrange dynamic model, India positions seventeenth among the nations at the most elevated hazard of importation of nCOVID-19 through air travel. The likelihood of a tainted air voyager to come to India as the last goal was 0.209 percent, with the most elevated relative import chance in Delhi (0.064%) followed by Mumbai, Kolkata, Bengaluru, Chennai, Hyderabad also, Kochi. This with regards to a plague that has effectively set in. In India, the initial 2019-nCoV disease case was accounted for on January 30, 2020, as indicated by the Ministry of Health and Family Welfare, Government of India. Huge numbers of the underlying cases in India originated from contact with the individuals having a background marked by going from Iran, Italy, and China. The first case was three people went from Wuhan, China to Kerala, India which is presently thought of as the focal point of the pandemic. Later all through India, the individuals who met voyagers from tainted nations contracted with the malady. First COVID-19 demise in the nation was accounted for on March 13^th^, 2020 (MOHFW, India). Although being a nation of 1.2 billion and a neighbour of China, India has genuinely contained the spread of COVID-19 contamination to date.

Currently, USA is worst affected with more than 456500 people infected of which more than 16000 died accounting to roughly about 3.6% mortality rate. The whole of Europe especially Italy, Spain, France and Germany are infected drastically. In the Indian subcontinent COVID-19 is still escalating and reached the milestone of 6725 infections and 226 deaths. Apart from India, 65 and 33 deaths were reported in Pakistan and Bangladesh respectively (World Health Organization 2020).

Mathematical model is very essential to study the outbreak of epidemics. Multiple projections can be generated by varying the exposure factor that influences then growth rate of infections. This is the major way out in order to provide insights for selective quarantining and lockdowns.

### SEIR Model

Kermack– McKendrick (1927) had first proposed the SEIR-type epidemiological models for the modelling of the Infectious disease studies. Infectious diseases are caused when certain types of parasites invade into a host. In SEIR models, people in a population are partitioned into susceptible (*S*), exposed/latent (*E*), infectious (*I*) and recovered/removed (*R*) individuals, and the models are alluded to on the premise of the contamination’s statuses of included people.

As a mediation, we demonstrated an ‘isolate of symptomatic’ situation wherein an extent p of symptomatic cases was isolated inside a normal of d long periods of creating indications. To fuse this mediation, we adjusted the model conditions as follows

Here,

Susceptible (S) people can be defined as the individuals who have a probability to get infected.

Exposed (E) people are those, who are not infectious, but they have been exposed to the disease and at the same time they are infected.

Infectious (I) people can spread the infection

Recovered (R) people are those, who have recovered from the infection

The evaluation and propagation of nCOVID-19 infections in India can also be modelled and predicted with the help of Susceptible-Exposed- Infectious- Recovered (SEIR) model, which is as follows,

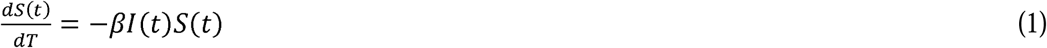

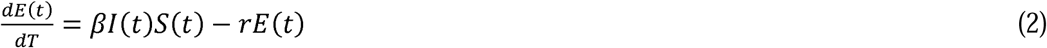

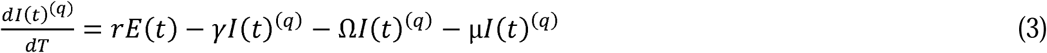

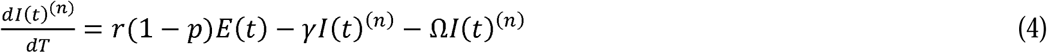

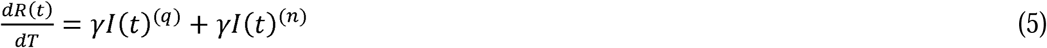

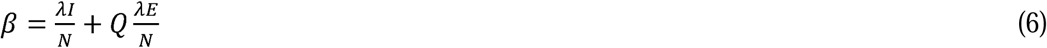

### Model States

*t* is the daily- time parameter

*S(t)* is the total susceptible population of India at time *t*

*E(t)* is the exposed population of India at time t

*I(t)* is the active infections of nCOVID-19 in India at time *t*

*R(t)* is the number of recoveries from nCOVID-19 in India at time t

*dS(t), d/(t), dR(t)* is the change in the state at time t.

### Mitigation

If nCOVID-19 began spreading in India, we built up a numerical model to recreate the transmission elements in India, just as their populace network. We picked to concentrate on these populace focuses on the supposition that the presentation of nCOVID-19 was well on the way to happen in worldwide transportation centre points, and consequently the urban areas were well on the way to be the central purposes of starting nCOVID-19 transmission in the nation.

### Model Parameter

The constant β signifies the growth rate, which is defined as the number of rises in infections of nCOVID-19 in India due to the interactions between the susceptible and infected populations. It depends on various factors such as population, reproduction number R_0_ of nCOVID-19, mobility, precautionary measures etc.

γ is defined as the rate of outcomes. In other words, it can be said as the rate at which the infected persons get recovered. It is assumed as the recovered persons will not spread the infection anymore, at least for the span of one month.

Ω is the per capita rate of death among symptomatic.

µ is the inverse of the average quarantine delay, d.

Q is the infectiousness of exposed cases, relative to symptomatic cases, is termed as relative infectiousness.

r is defined as, per capita rate of developing symptoms among those exposed.

λ is the average number of infections caused per day per symptomatic case.

P_total_ is the total population of India as per latest available data. C_0_ and S_0_ are the initial number of Susceptible individuals.

### Parameters Based on Indian Trends

Contact rates are defined as the number of susceptible people, an infectious person contact. Among 7,290 individuals in eight European nations, the normal contacts per individual was 13.4 with a nation scope of 7.95 (Germany) to 19.77 (Italy). Young people had the most noteworthy contact rate (18) contrasted with adults 20-60 (13), and olds 60+ (8) ^[7-8]^. For the case of India, we have assumed a contact rate of 12 to optimize the model.

Transmissibility is defined as the probability that a contact between a susceptible person and an infected person results in infection. The value used, 1.7%, was estimated by comparing the Reproduction Number (R_0_) in community settings (1.4 to 3.9) with a normal contact rate and a cruise ship (14.8) which has a high contact rate. R_0_ varies with contact rate whereas transmissibility and removal rate (inverse of duration of infectiousness) were kept constant.^[9-10].^

Reproduction Number (R_0_) is defined as the number of secondary cases resulting from one case. Mathematically it can be obtained as the product of Contact Rate, Transmissibility and Removal Rate. Estimates for nCOVID-19 have been between 1.4 to 3.9. The inverse of R_0_, (1-(1/R_0_)) is the proportion of the population who need to be infected (or vaccinated) for transmission to no longer be self-sustaining. R_0_ is also an indication of the effectiveness of community interventions. An R_0_ less than 1 indicates transmission has stopped ^[11]^.

The authors have found that from 10 early cases in China showed the mean serial interval (time between successive cases) was 7.5 days with a Standard Deviation (SD) of 3.4 days. A more recent estimate among 468 cases was 3.96 days with a 4.75-day SD ^[12-13]^. But in the current context of India, the authors noticed that, a patient is taking approximately 14 days to get recovered. So, for this case of nCOVID-19 modelling of India, the authors have taken duration of Infectiousness as 14 days.

Various errors such as square integral error, terminal error and terminal rate error between the actual data and predicted data are minimized using optimizing different parameters ^[14]^.

### Simulation Results

After 15th of March. 2020, nCOVID-19 infection in India started to show an exponential trend. The SEIR model has been simulated and all the parameters have been optimised based on infection trends obtained for India for nearly 20 days (up to 6th of April 2020). The differential equations have been solved by using Python and R-Programming languages. The forecasted data are showed graphically in Figure 1.

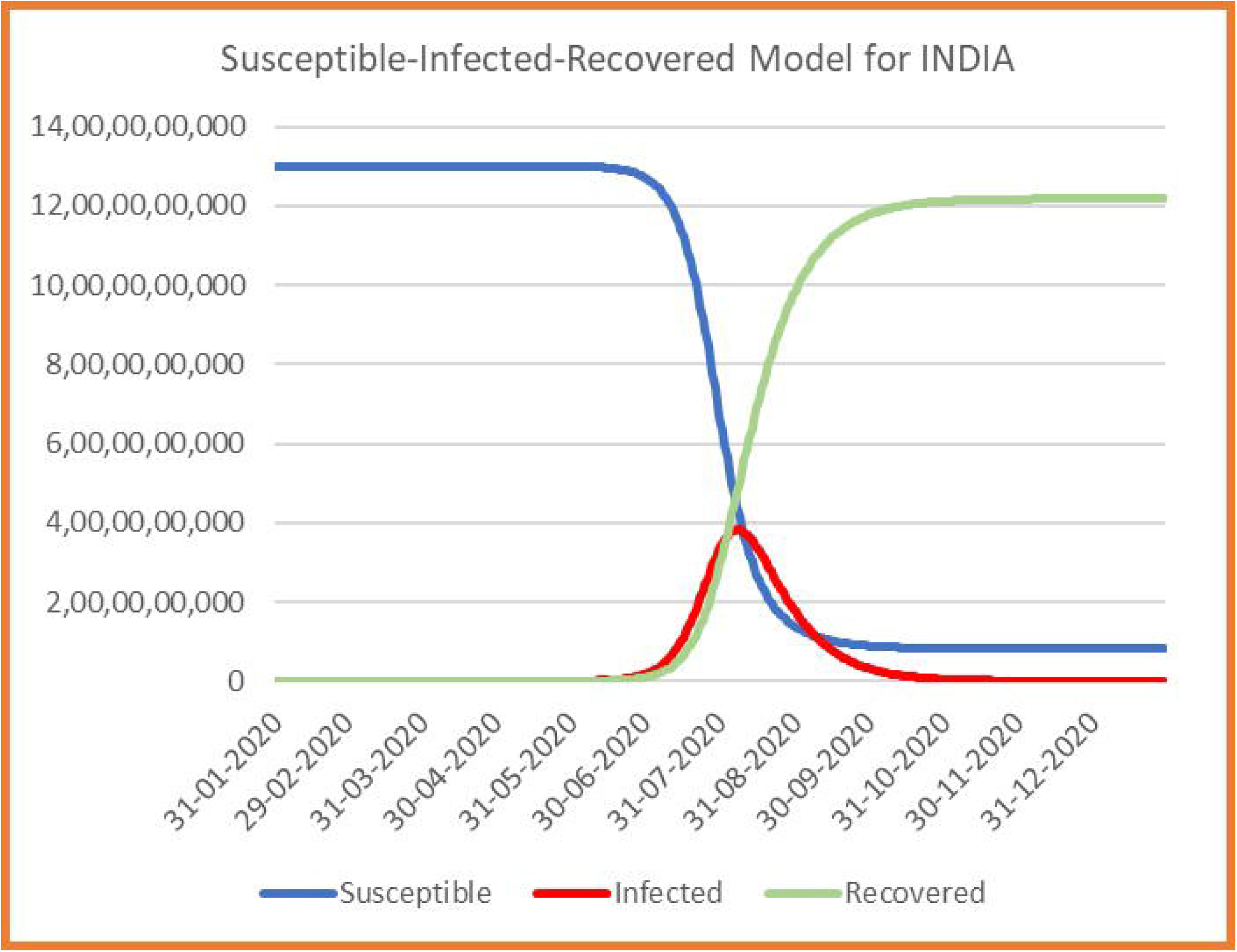

From Figure 2, it is clearly evident that the number of forecasted infected patients are really correlating with actual number of infected patients till date. Here the validity and novelty of the model is established. So, this model can be used to predict the future scenario of nCOVID-19 in India. The authorss outlined how these effects may shift, under various transmission and mediation situations.

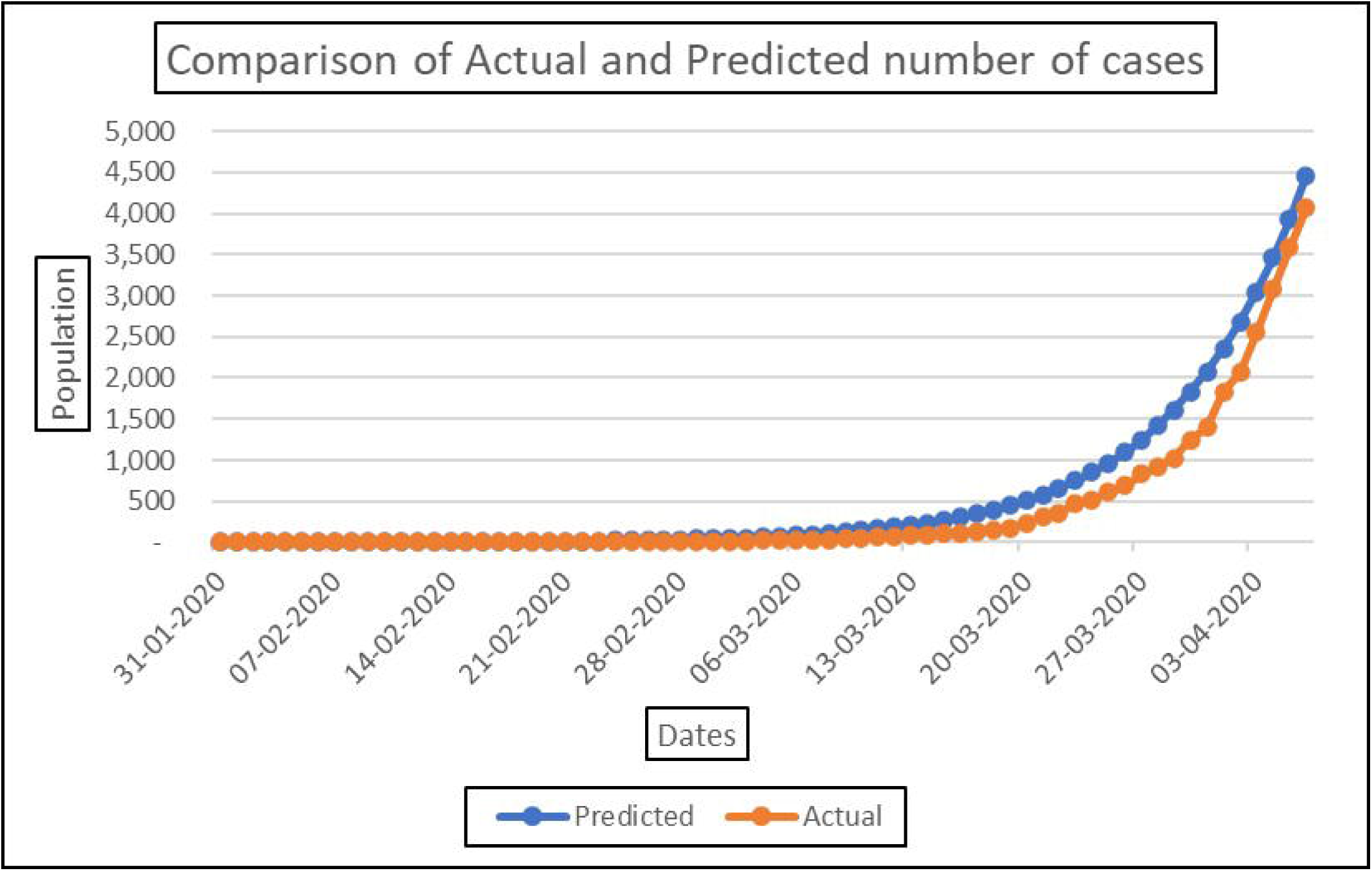

### Nationwide projections for India

By solving the course of equations, the authors have obtained the trends of Corona virus infected persons in India since 31^st^ of January 2020, the first reporting of nCOVID-19 case in India. The authors have calculated the number of infected and recovered people on daily basis. This model has showed in Figure 2, a very good correlation with the original data of number of infected individuals.

From the above obtained Figure 2, the authors have concluded that the developed model stands very good in the current scenario of nCOVID-19 infection in India.

But the alarming fact is that, depending upon the future predictions by the model, India in a great danger zone. As If this trend is going on, then within a nearby future the number of nCOVID-19 infections will grow exponentially and will be out of control. This prediction can be showed with the help of Figure 3.

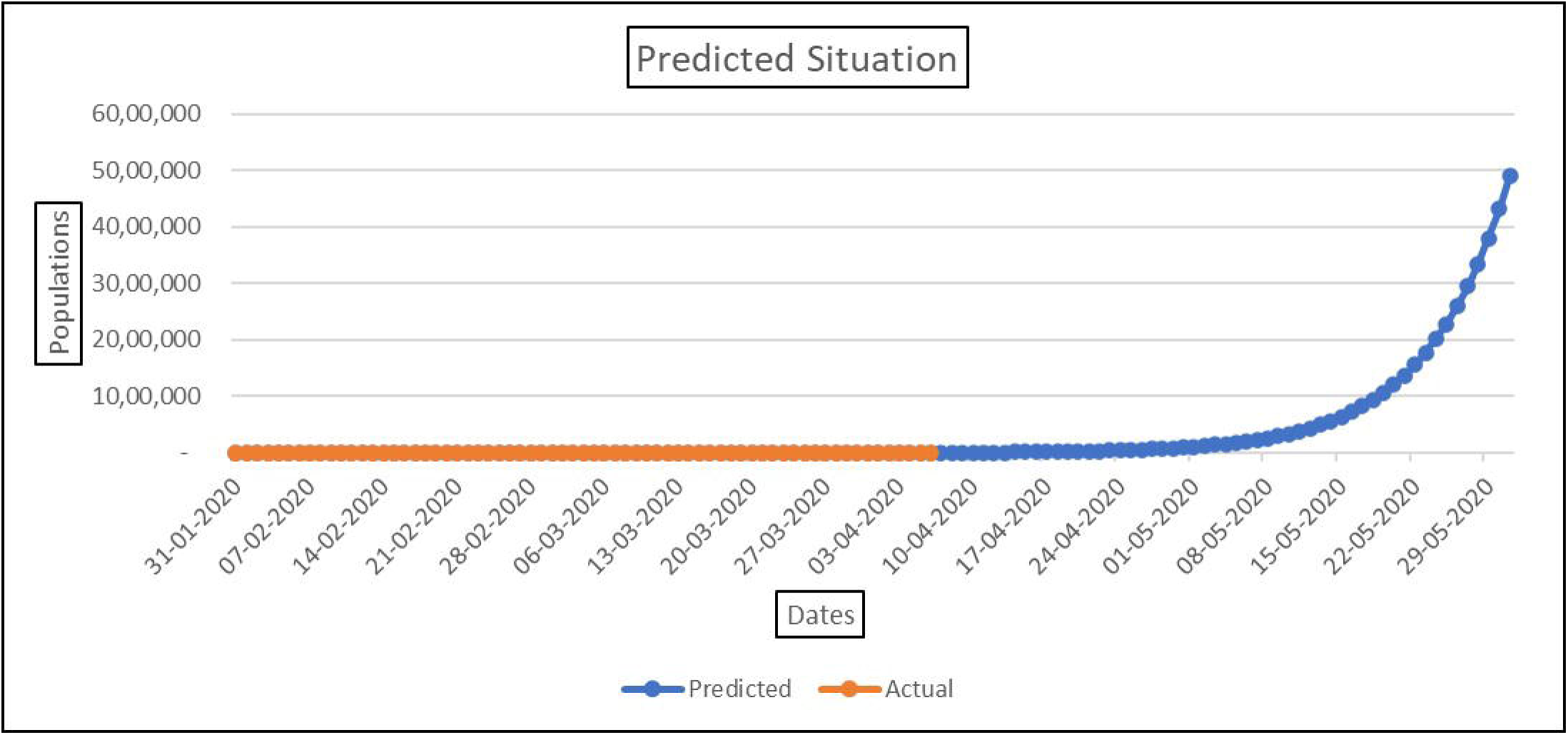

If this curve in Figure 3, goes on increasing by following the similar trends, then the model predicts that the total number of nCOVID-19 infected individuals will reach the milestone of approximately 50,00,000 within the month of June 2020. Then the situation of India will be worst and can not be controlled by any means. It will destroy the whole socio-economic balance of India. Current Mortality rate till 08.03.2020 is nearly about 3%. If this mortality rate persists then a dangerous situation will happen in India. As the development pace of COVID-19 contaminations ascended on 21st March 2020, the Prime Minister of India Shri. Narendra Modi announced a supposed one-day lockdown on 22nd March 2020 in name ‘Janata Curfew’ which brought down the development pace of cases from 3.69 to 1. (MOHFW, India). This brought about the statement of a multi-day long further lockdown in the nation to fourteenth April 2020. On the fifth day of the lockdown again an ascent in development pace of contaminated people was watched. This brought up the issue of whether the lockdown is powerful or not. Being a nation of 100 million individuals the viability of conferring lockdown is a significant obstacle to the Government. A minor level of individuals despite everything act slack and careless about the lockdown have become super-spreaders of their locale. Indeed, even 0.1% of oblivious individuals of the nation are an immense populace that can spread the pandemic vivaciously. Another explanation behind an up ascend in new cases during the lockdown time frame might be because of wasteful screening rehearses. Shortage of volunteers, inaccessibility of screening units, inappropriate wilful detailing of patients may likewise be a portion of the fundamental driver of wasteful screening and slanted insights. Be that as it may, the lockdown has significantly hindered the contamination rate contrasted with different nations. It is a way to control this pandemic situation by maintain social distances. But as decided by Indian Government if, after 14^th^ of April 2020, the total lock down will be relaxed in India, it will increase the factors like contact rate, transmissibility and reproduction number.

So, to control this situation, the authors are suggesting some points by simulating the SEIR model

- If the total lock down will be relaxed, then it must be implemented again after a 5 days relaxation.
- The main parameter which affects the total number of infected individuals is the contact rate. If a normal patient, by any chance goes near the vicinity of a nCOVID-19 patient without any preventive measurements, then the probability of getting infected of a susceptible people increase rapidly. So, to overcome this situation, the contact rate should be taken care.
- If the contact rate can be maintained within a range of 5, then the chance to control this pandemic situation get increased.
- The transmissibly is another major factor, which can control the situation. If the nation can maintain the probability of a susceptible people to get infected, below 10%, then it will help to overcome the pandemic in the context of India.

Such measures could reduce the peak prevalence substantially, thus minimizing the pressure on public health services. It is clearly seen that the peak of the graph in Figure 4 is lowered with a considerable extent by implementing the mentioned conditions. The total number of nCOVID-19 cases are also decreasing day by day and the reproduction number (R_0_) reaches below 1. It is clearly visible that the number of recovered people is increasing, and the infection is vanishing. The comparison of the simulated SEIR model with the modified SEIR model is shown in Figure 5. Therefore, the lock-down has the effect of ‘flattening’ the epidemic curve, distributing cases over a longer duration than in the absence of lock-down. The intervention could reduce the cumulative incidence by nearly 60 per cent.

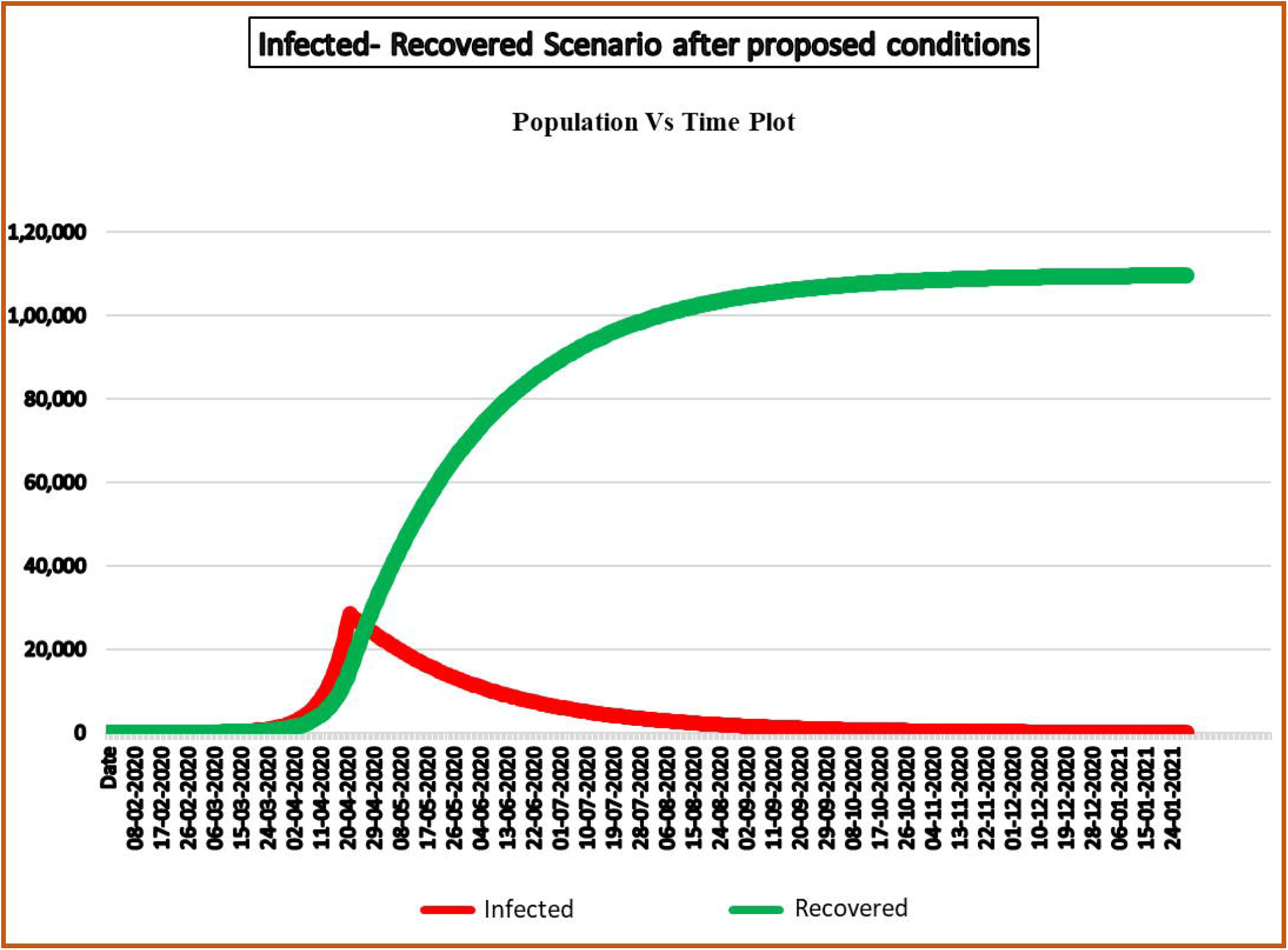

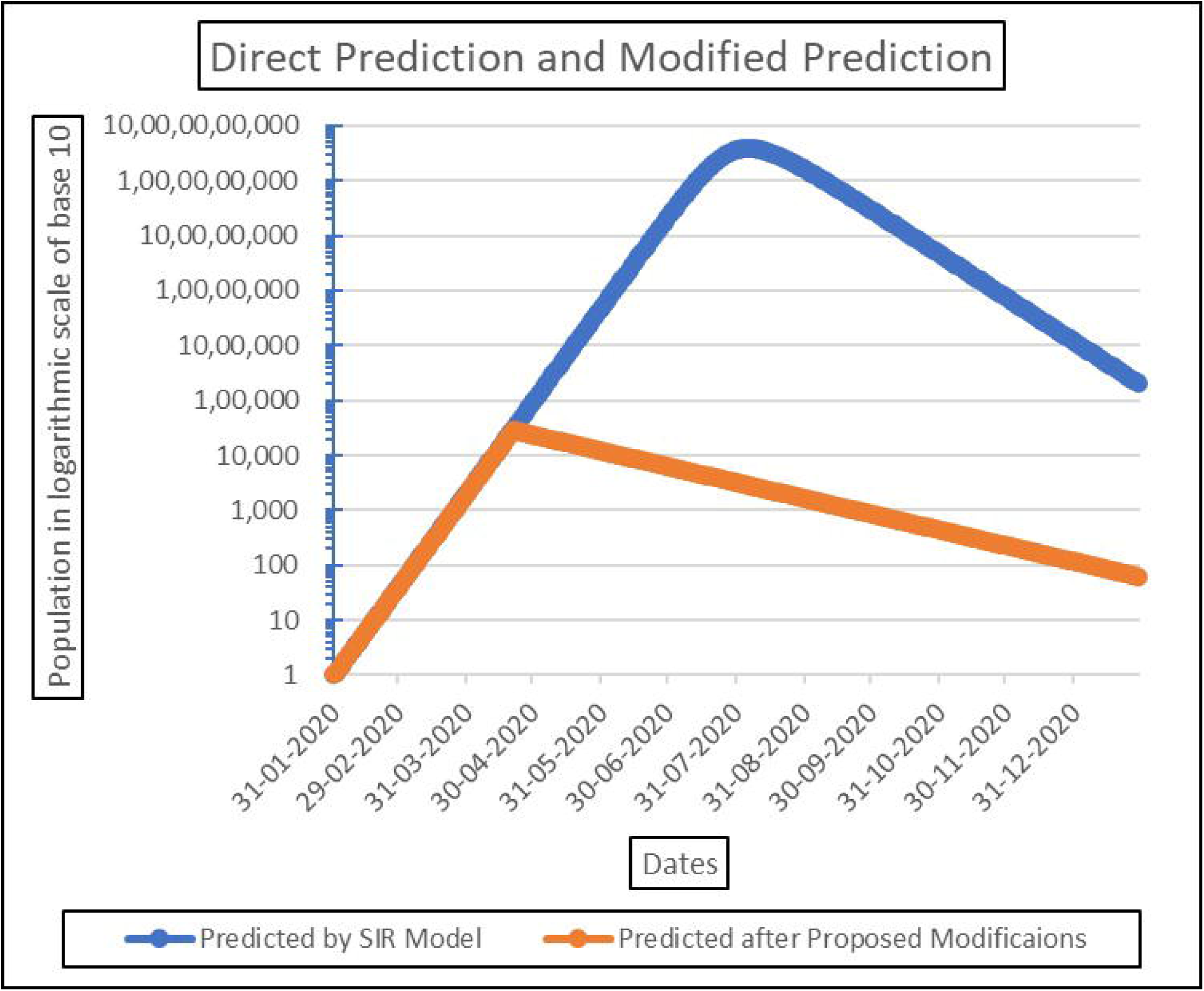

## Conclusions

From the mentioned SEIR model, the authors have simulated the current context of nCOVID-19 cases of India. They provided a clear idea about the growths and trend of the pandemic in India. They also showed the data, if the lockdown gets relaxed, then the outburst of infection will occur in India. So, to overcome that situation, the authors are suggesting that the implementation lockdown again for 21-28 days is very necessary. As it will keep the social distancing as well as the lockdown will influence the above-mentioned parameters to flatten the peak of infection curve and decrease the number of infection cases. This is a persistent work where the authors are attempting to discover the model parameters regular and venture the potential situations, by fluctuating the exposure factor for the pace of contamination, as an aftereffect of developing degrees of isolating. While this isn’t a totally unquestionable projection, the model parameters look very reliable, anyway they may mirror an over estimation of the anticipated number of contaminations to redress for the unreported or undetected contaminations. In this manner, we may take the numbers right now be a guide for additional activity by the law upholding specialists.

## Data Availability

All data are collected from World Health Organisation's website and MOHFW website of India

https://www.mohfw.gov.in/

